# Clinical and Laboratory Profiles of 75 Hospitalized Patients with Novel Coronavirus Disease 2019 in Hefei, China

**DOI:** 10.1101/2020.03.01.20029785

**Authors:** Zonghao Zhao, Jiajia Xie, Ming Yin, Yun Yang, Hongliang He, Tengchuan Jin, Wenting Li, Xiaowu Zhu, Jing Xu, Changcheng Zhao, Lei Li, Yi Li, Hylemariam Mihiretie Mengist, Ayesha Zahid, Ziqin Yao, Chengchao Ding, Yingjie Qi, Yong Gao, Xiaoling Ma

## Abstract

The outbreak of the novel coronavirus disease 2019 (COVID-19) infection began in December 2019 in Wuhan, and rapidly spread to many provinces in China. The number of cases has increased markedly in Anhui, but information on the clinical characteristics of patients is limited. We reported 75 patients with COVID-19 in the First Affiliated Hospital of USTC from Jan 21 to Feb 16, 2020, Hefei, Anhui Province, China. COVID-19 infection was confirmed by real-time RT-PCR of respiratory nasopharyngeal swab samples. Epidemiological, clinical and laboratory data were collected and analyzed. Of the 75 patients with COVID-19, 61 (81.33%) had a direct or indirect exposure history to Wuhan. Common symptoms at onset included fever (66 [88.0%] of 75 patients) and dry cough (62 [82.67%]). Of the patients without fever, cough could be the only or primary symptom. The most prominent laboratory abnormalities were lymphopenia, decreased percentage of lymphocytes (LYM%), decreased CD4^+^ and CD8^+^ T cell counts, elevated C-reactive protein (CRP) and lactate dehydrogenase (LDH). Patients with elevated interleukin 6 (IL-6) showed significant decreases in the LYM%, CD4^+^ and CD8^+^ T cell counts. Besides, the percentage of neutrophils, CRP, LDH and Procalcitonin levels increased significantly. We concluded that COVID-19 could cause different degrees of hematological abnormalities and damage of internal organs. Hematological profiles including LYM, LDH, CRP and IL-6 could be indicators of diseases severity and evaluation of treatment effectiveness. Antiviral treatment requires a comprehensive and supportive approach. Further targeted therapy should be determined based on individual clinical manifestations and laboratory indicators.

## Introduction

Since Dec 2019, a series of acute respiratory illness outbreaks in Wuhan, Hubei Province, China [1, 2]. The disease has been subsequently identified in other provinces in China, and other counties. On Jan 7, a novel coronavirus was identified by deep sequencing analysis of samples from throat swabs and lower respiratory tract. The disease caused by the novel virus is now named by WHO as novel coronavirus disease 2019 (COVID-19). Epidemiological research shows that all infected patients had travel or residence records in Wuhan, suggesting the possibility of person-to-person transmission [3]. By Feb 22, 2020, more than 75,000 confirmed cases, including 1716 health-care workers, have been identified in China. And 989 patients have been diagnosed in Anhui Province, including 6 deaths.

The novel coronaviruse is an enveloped non-segmented positive sense RNA virus belonging to the betacoronaviruses. The well-known atypical pneumonia virus (SARS-CoV) and Middle East Respiratory Syndrome Virus (MERS-CoV) are also betacoronaviruses [4]. Clinical manifestations of COVID-19 include fever, dry cough, myalgia and fatigue. Symptoms of headache, expectoration, and diarrhea seem to less common. Radiographic evidence suggested pneumonia. About half of patients have developed severe pneumonia. Nearly one third of patients require intensive care because of acute respiratory distress syndrome (ARDS) or multiple organ failure [1, 5].

At present, there are relatively few reports about novel coronavirus pneumonia in Anhui Province. Here, we described the epidemiological, clinical and laboratory characteristics of 75 COVID-19 confirmed patients admitted to the First Affiliated Hospital of USTC, Hefei. This study will be beneficial for the diagnosis and treatment of COVID-19 patients in clinical practice.

## Methods

### Patients

In this study, we eventually enrolled 75 patients from the First Affiliated Hospital of USTC between Jan 21, and Feb 16, 2020. Most patients came to the hospital because of fever or respiratory symptoms. Our clinical team consulted and recorded their epidemiological history in detail regarding to whether they had been to Wuhan or exposed to people who came from Wuhan recently. Nasopharyngeal and throat swabs were taken for respiratory pathogens test. The physical findings, hematological, biochemical and radiological results were also recorded. All patients were identified as laboratory-confirmed COVID-19 infection. All patients enrolled in this study were diagnosed according to World Health Organization interim guidance. The study was approved by the Ethics Committee of the First Affiliated Hospital of USTC.

### Procedures

Respiratory nasopharyngeal swabs were collected and the presence of COVID-19 was detected by next real-time RT-PCR methods. Viral RNA was extracted using QIAamp RNA virus Kit (Qiagen, Heiden, Germany). The diagnostic test was done using a commercial coronavirus test kit (Shenzhen Huada Yinyuan Pharmaceutical Technology Co., Ltd., Shenzhen). The specific primers and probe targeted to nucleocapsidprotein (N) were used and the sequences were as follows: forward primer 5′-GGGGAACTTCTCCTGCTAGAAT-3′; reverse primer 5′-CAGACATTTTGCTCTCAAGCTG-3′; and the probe 5’-FAM-TTGCTGCTGCTTGACAGATT-TAMRA-3’. Conditions for the amplifications were 50°C for 20 min, 95°C for 10 min, followed by 40 cycles of denaturation at 95°C for 15 s and extending and collecting fluorescence signal at 60°C for 30 s. A cycle threshold value (Ct-value) less than 37 was defined as a positive test result, and a Ct-value of 40 or more was defined as a negative test. A medium load, defined as a Ct-value of 37 to less than 40, requires a retesting according to the guideline of Chinese Centers for Disease Control and Prevention (http://ivdc.chinacdc.cn/kyjz/202001/t20200121_211337.html).

We also examined other respiratory viruses, including influenza, avian influenza, respiratory syncytial virus, adenovirus, parainfluenza virus, SARS-CoV and MERS-CoV, with realtime RT-PCR. Hematological parameters including blood routine, blood biochemistry, coagulation profile, and infection-related biomarkers were recorded. Plasma cytokine interleukin 6 (IL-6) levels were detected by ELISA. And the CD4^+^ and CD8^+^ T cell subsets were counted using flow cytometry.

### Statistical analysis

We presented continuous measurements as median (IQR) and categorical variables as number (%). Continuous variables were analyzed using the Mann-Whitney test. For laboratory results, we also assessed whether the measurements were outside the normal range. Graphpad prism 8.3 was used for all analyses. A two-sided α of less than 0.05 was considered statistically significant.

## Results

Totally, 75 patients diagnosed with COVID-19 were included in this study. Among them, 61 (81.33%) patients had been to Wuhan or exposed to people who came from Wuhan. The median age of the patients was 47 years. Among them, 36 (48%) were aged 40-59 years, 25 (33.3 %) were aged 20-39 years, 11 (14.67%) were aged 60-79 years. The youngest patient aged 16 years and the oldest aged 91 years. More than half of the participants were men (42 [56%]). Twenty-nine (38.67%) patients had one or more chronic diseases, including cardiovascular and cerebrovascular disease, diabetes, chronic kidney disease, respiratory system disease, nervous system disease, chronic liver diseases, and malignant tumor (Table 1).

**Table 1.** Demographics and baseline characteristics of 75 patients infected with COVID-19.

| Characteristics | No. (%) |
| --- | --- |
| Age, years, Median (IQR) | 47 (34-55) |
| Range | 16-91 |
| <20 | 1 (1.33%) |
| 20-39 | 25 (33.33%) |
| 40-59 | 36 (48.00%) |
| 60-79 | 11 (14.67%) |
| ≥80 | 2 (2.67%) |
| Sex |  |
| Female | 33 (44%) |
| Male | 42 (56%) |
| Exposure to Wuhan people | 61 (81.33%) |
| Chronic medical illness | 29 (38.67%) |
| Cardiovascular and cerebrovascular diseases | 16 (21.33%) |
| Diabetes | 6 (8.00%) |
| Chronic kidney disease | 4 (5.33%) |
| Chronic liver disease | 4 (5.33%) |
| Respiratory system disease | 2 (2.67%) |
| Nervous system disease | 1 (1.33%) |
| Malignant tumour | 1 (1.33%) |

Abbreviations: COVID-19, coronavirus disease 2019; IQR, interquartile range.
Data are presented as median (IQR) or n/N (%). N is the total number of patients with available data.

Most patients admitted to hospital because of fever (66 [88.0%]) and dry cough (62 [82.67%]). Nearly a third of patients had chest tightness (24 [32.0%]). And 20 (26.67%) patients had all the three symptoms mentioned above. Less common symptoms included sputum production (22 [29.33%]), fatigue (17 [22.67%]), muscle soreness (9 [12.0%]) and poor appetite (9 [12.0%]). Other symptoms included diarrhea, sore throat, headache, shortness of breath and stomach ache. Nine patients had a body temperature below 37.3°C, and all of them had symptom of dry cough. Only a small proportion had sputum, fatigue, poor appetite and chest tightness (Table 2).

**Table 2.**
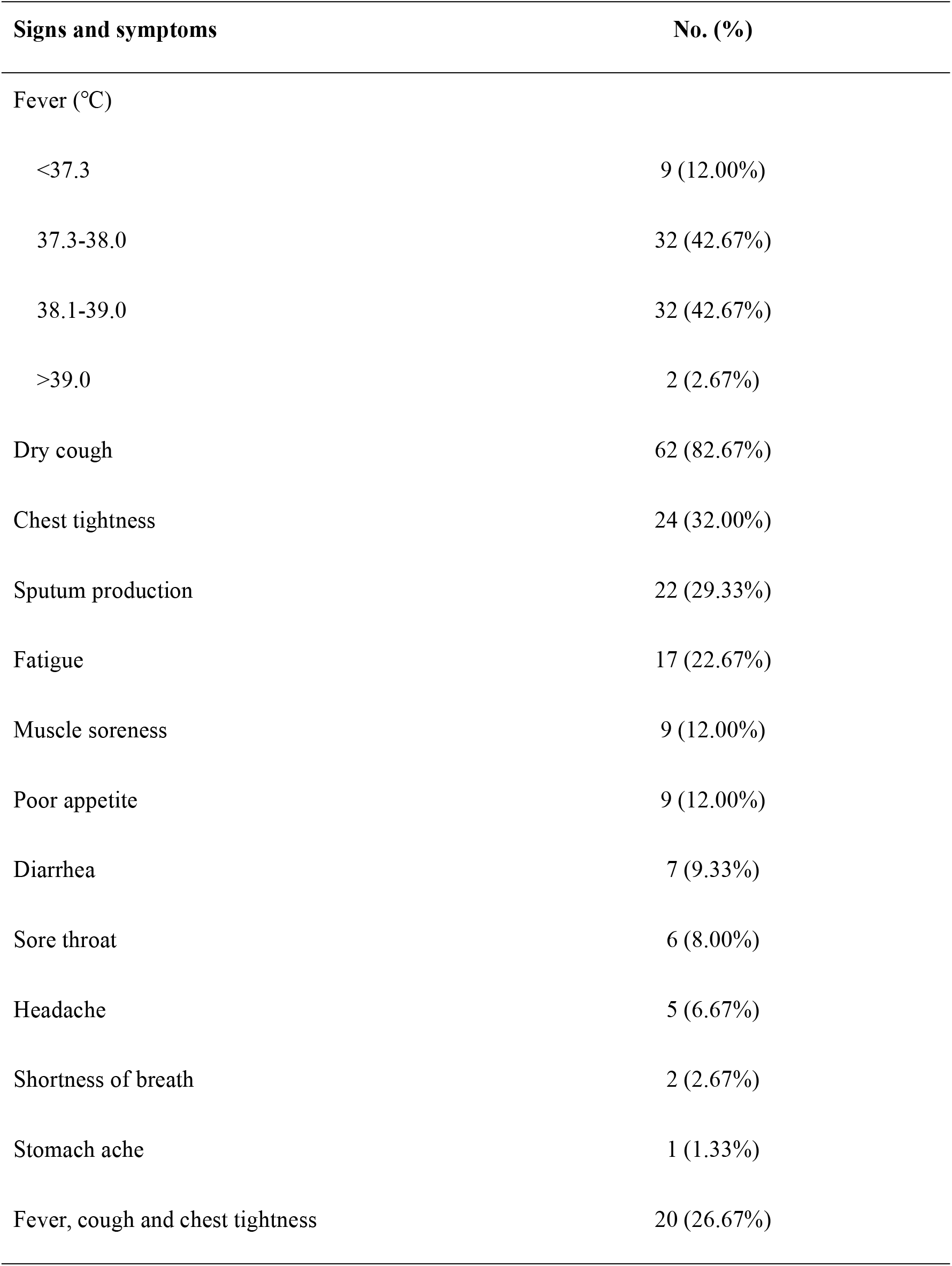
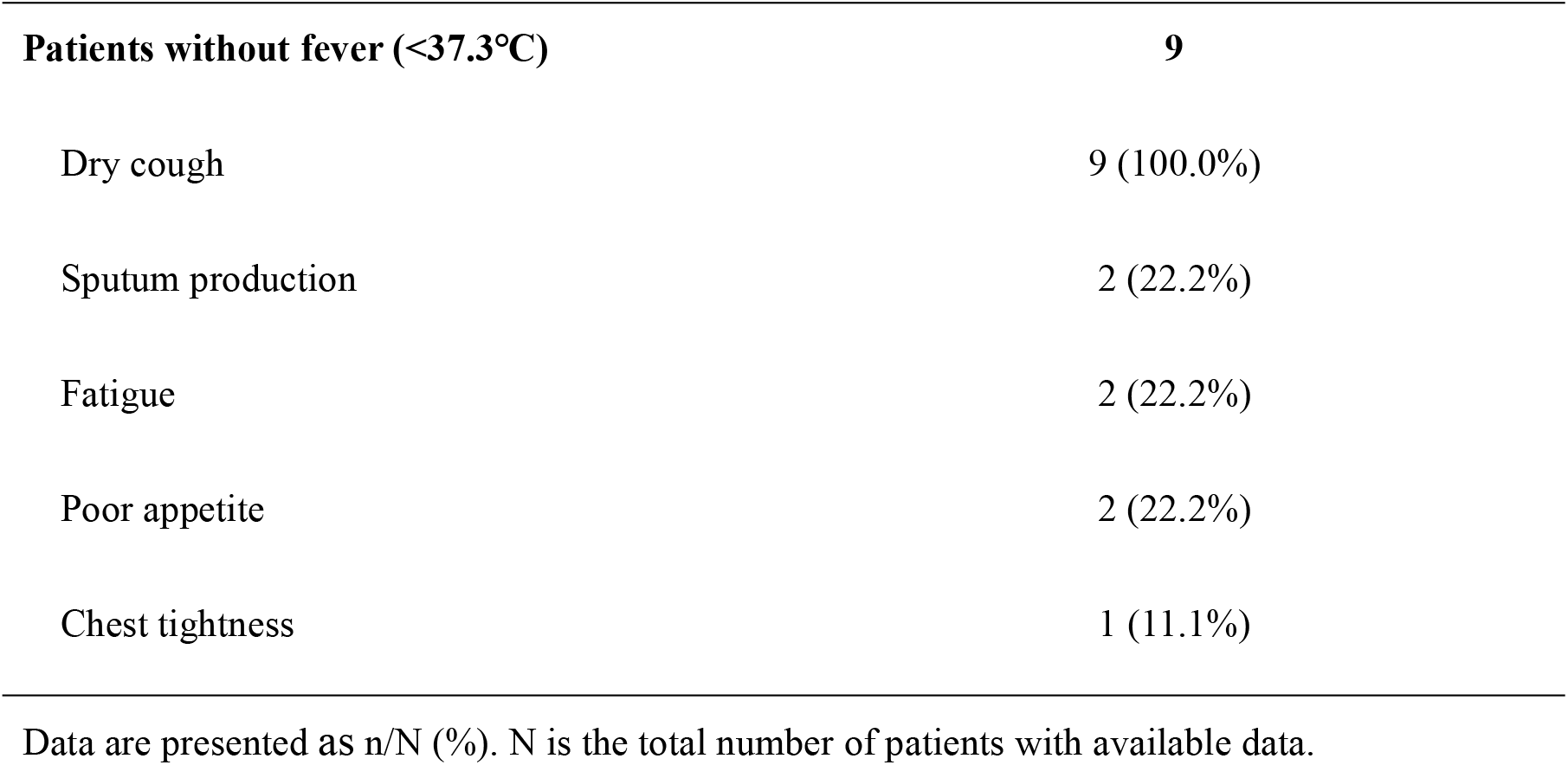
Signs and symptoms of patients with COVID-19.

| Signs and symptoms | No. (%) |
| --- | --- |
| Fever (°C) |  |
| <37.3 | 9 (12.00%) |
| 37.3-38.0 | 32 (42.67%) |
| 38.1-39.0 | 32 (42.67%) |
| >39.0 | 2 (2.67%) |
| Dry cough | 62 (82.67%) |
| Chest tightness | 24 (32.00%) |
| Sputum production | 22 (29.33%) |
| Fatigue | 17 (22.67%) |
| Muscle soreness | 9 (12.00%) |
| Poor appetite | 9 (12.00%) |
| Diarrhea | 7 (9.33%) |
| Sore throat | 6 (8.00%) |
| Headache | 5 (6.67%) |
| Shortness of breath | 2 (2.67%) |
| Stomach ache | 1 (1.33%) |
| Fever, cough and chest tightness | 20 (26.67%) |
| **Patients without fever (<37.3°C)** | **9** |
| Dry cough | 9 (100.0%) |
| Sputum production | 2 (22.2%) |
| Fatigue | 2 (22.2%) |
| Poor appetite | 2 (22.2%) |
| Chest tightness | 1 (11.1%) |

Data are presented as n/N (%). N is the total number of patients with available data.

The blood counts of patients on admission showed leucopenia (white blood cell counts below the normal range; 12 [16.0%]). Twenty-nine (38.67%) patients showed increased neutrophil percentage (NEU%). Over half of the patients (40 [53.33%]) showed lymphopenia (lymphocytes counts less than 1.1×10^9^/L). However, no patients had increased lymphocytes counts. Thirty-one (41.33%) and 28 (37.33%) patients showed decreased counts of CD4^+^ and CD8^+^ T cell levels, respectively. The CD4^+^/CD8^+^ ratio was below the normal range in 11 (14.67%) patients. Haemoglobin were decreased in 11 (14.67%) patients and increased in 18 (24%) patients. Platelets were below the normal range in 14 (18.67%)) patients and above the normal range in only 2 (2.67%) patients. Most patients showed impaired coagulation function. Activated partial thromboplastin time (APTT) was longer in 44 (58.67%) patients and prothrombin time (PT) was longer in 30 (40%) patients (Table 3).

**Table 3.**
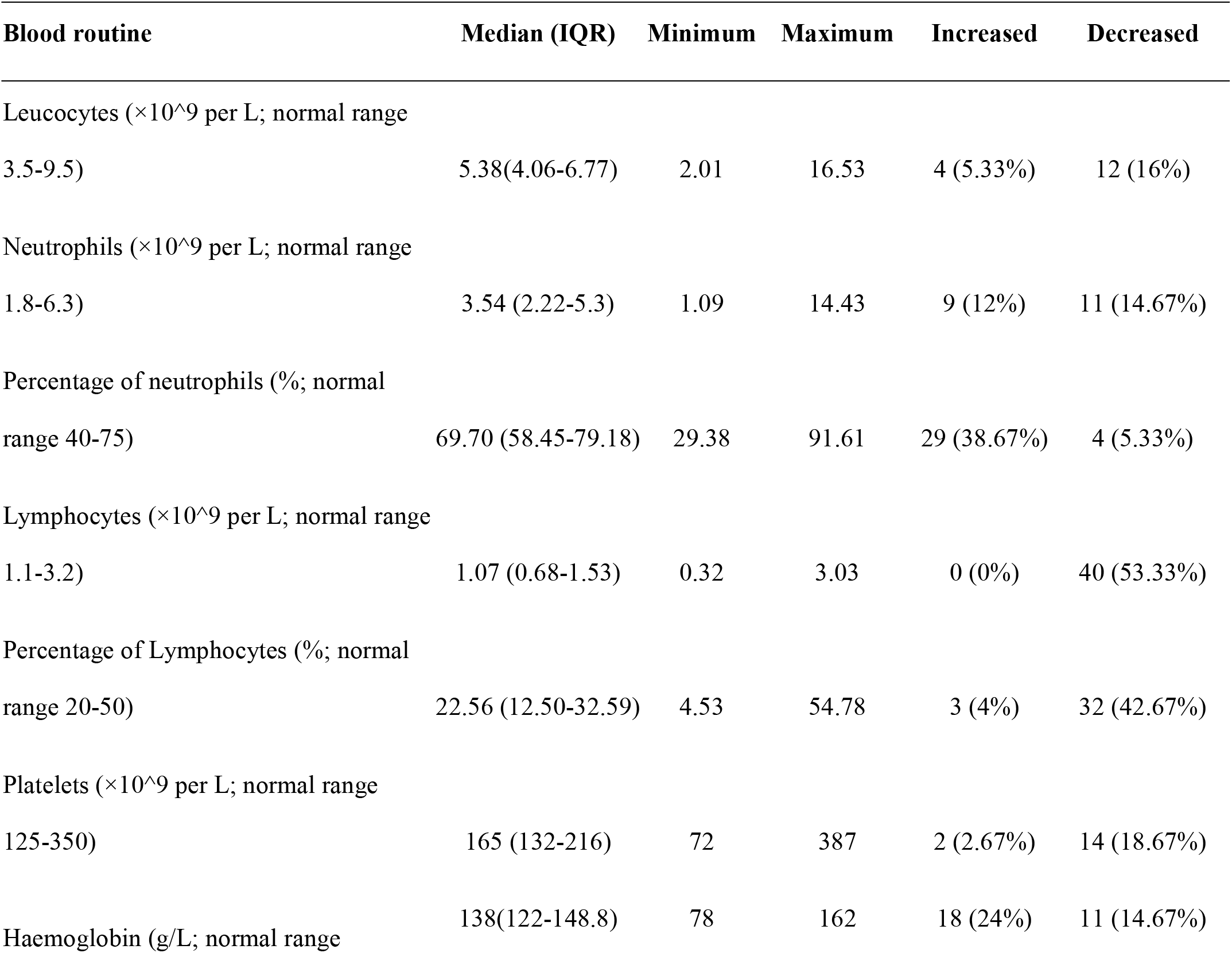
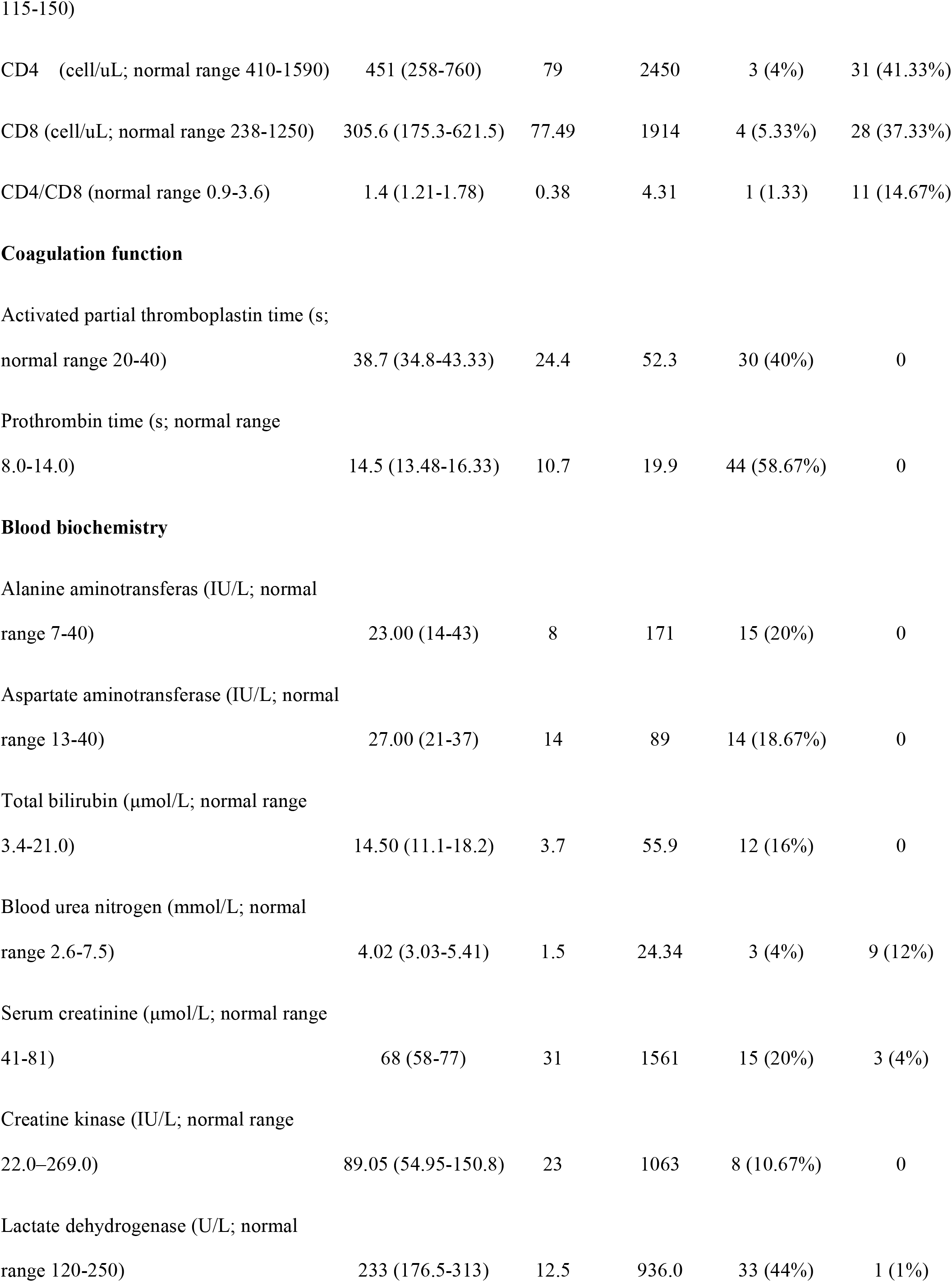
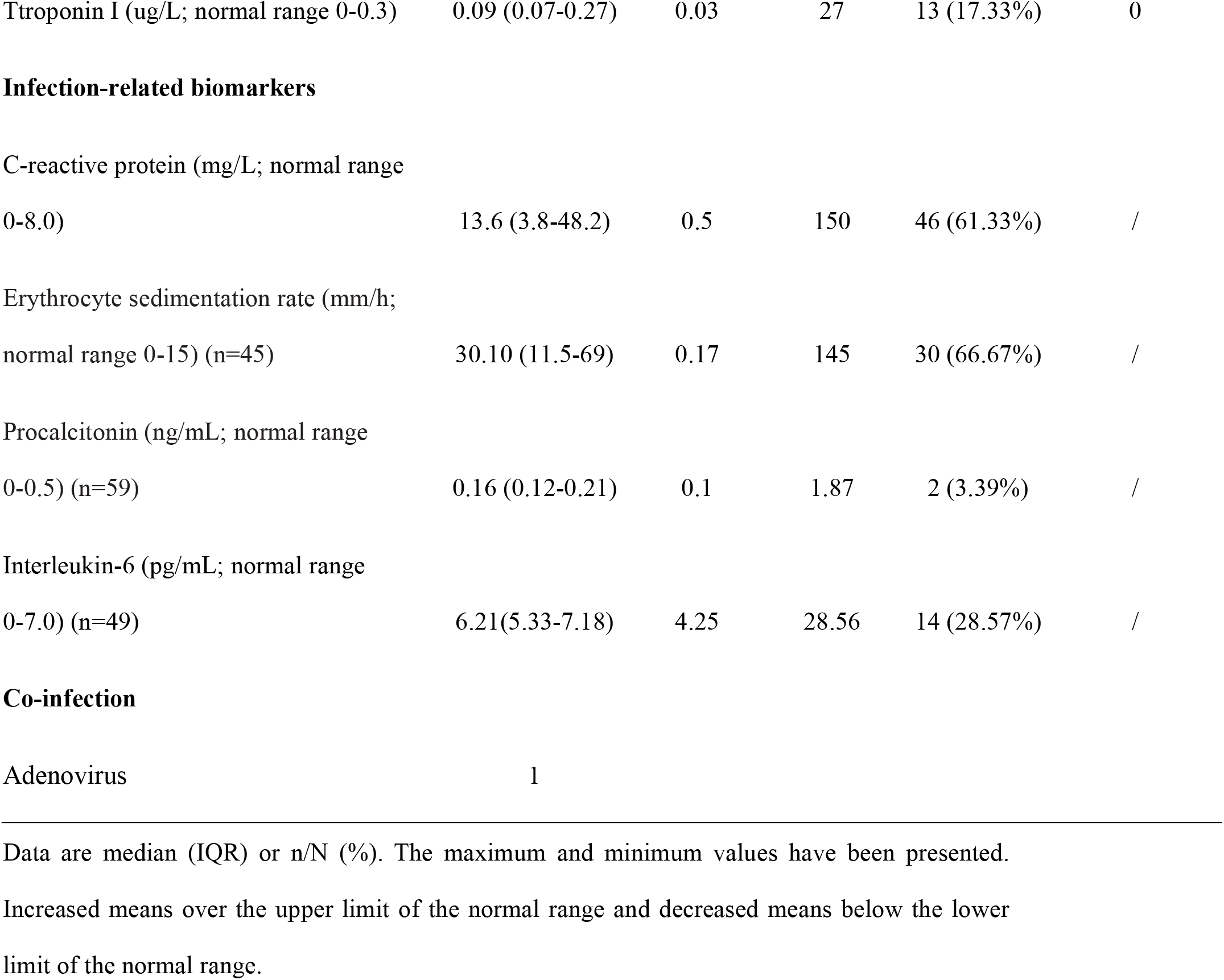
Laboratory results of patients infected with COVID-19 on admission to hospital.

| **Blood routine** | **Median (IQR)** | **Minimum** | **Maximum** | **Increased** | **Decreased** |
| --- | --- | --- | --- | --- | --- |
| Leucocytes (×10<sup>9</sup> per L; normal range 3.5-9.5) | 5.38(4.06-6.77) | 2.01 | 16.53 | 4 (5.33%) | 12 (16%) |
| Neutrophils (×10<sup>9</sup> per L; normal range 1.8-6.3) | 3.54 (2.22-5.3) | 1.09 | 14.43 | 9 (12%) | 11 (14.67%) |
| Percentage of neutrophils (%; normal range 40-75) | 69.70 (58.45-79.18) | 29.38 | 91.61 | 29 (38.67%) | 4 (5.33%) |
| Lymphocytes (×10<sup>9</sup> per L; normal range 1.1-3.2) | 1.07 (0.68-1.53) | 0.32 | 3.03 | 0 (0%) | 40 (53.33%) |
| Percentage of Lymphocytes (%; normal range 20-50) | 22.56 (12.50-32.59) | 4.53 | 54.78 | 3 (4%) | 32 (42.67%) |
| Platelets (×10<sup>9</sup> per L; normal range 125-350) | 165 (132-216) | 72 | 387 | 2 (2.67%) | 14 (18.67%) |
| Haemoglobin (g/L; normal range | 138(122-148.8) | 78 | 162 | 18 (24%) | 11 (14.67%) |
| CD4 (cell/uL; normal range 410-1590) | 451 (258-760) | 79 | 2450 | 3 (4%) | 31 (41.33%) |
| CD8 (cell/uL; normal range 238-1250) | 305.6 (175.3-621.5) | 77.49 | 1914 | 4 (5.33%) | 28 (37.33%) |
| CD4/CD8 (normal range 0.9-3.6) | 1.4 (1.21-1.78) | 0.38 | 4.31 | 1 (1.33) | 11 (14.67%) |
| Troponin I (ug/L; normal range 0-0.3) | 0.09 (0.07-0.27) | 0.03 | 27 | 13 (17.33%) | 0 |
| Infection-related biomarkers |  |  |  |  |  |
| C-reactive protein (mg/L; normal range 0-8.0) | 13.6 (3.8-48.2) | 0.5 | 150 | 46 (61.33%) | / |
| Erythrocyte sedimentation rate (mm/h; normal range 0-15) (n=45) | 30.10 (11.5-69) | 0.17 | 145 | 30 (66.67%) | / |
| Procalcitonin (ng/mL; normal range 0-0.5) (n=59) | 0.16 (0.12-0.21) | 0.1 | 1.87 | 2 (3.39%) | / |
| Interleukin-6 (pg/mL; normal range 0-7.0) (n=49) | 6.21(5.33-7.18) | 4.25 | 28.56 | 14 (28.57%) | / |

Co-infection
|  |  |
| --- | --- |
| Adenovirus | 1 |

Data are median (IQR) or n/N (%). The maximum and minimum values have been presented.
Increased means over the upper limit of the normal range and decreased means below the lower limit of the normal range.

Fifteen patients had differing degrees of liver function abnormality, with alanine aminotransferase (ALT) or aspartate aminotransferase (AST) above the normal range. One patient with no underlying disease had a serious liver function damage (ALT 171 U/L, AST 60 U/L). Nearly half of patients showed abnormal myocardial zymogram, with the elevation of lactate dehydrogenase (LDH) in 33 (44%) patients and the elevation of Troponin I in 13 (17.33%) patients. Fifteen (20%) patients had different degrees of renal function damage with elevated serum creatinine. One patient with uremia had creatinine level of 1561 μmol/L (Table 3). These findings suggested that the internal organs could also be potential targets of COVID-19.

Regarding the infection index, most patients showed elevated C-reactive protein (CRP) and Erythrocyte sedimentation rate (ESR) levels. Procalcitonin (PCT) was elevated in 2 out of 59 patients. Forty-nine patients were tested for IL-6, and 14 (28.57%) of them showed levels above the normal range (Table 3). Further analysis showed that the 14 patients had significant decreases in lymphocytes percentage, CD4^+^ and CD8^+^ T cell counts, compared to those with normal IL-6 range. Besides, the NEU%, CRP and LDH levels increased significantly (Table 4; Figure 1). PCT values were within normal range in both two groups. These data indicated that there might be correlation between the increased IL-6 level and the severity of viral infection. And we will continue paying attention to this point in the future.

**Table 4.**
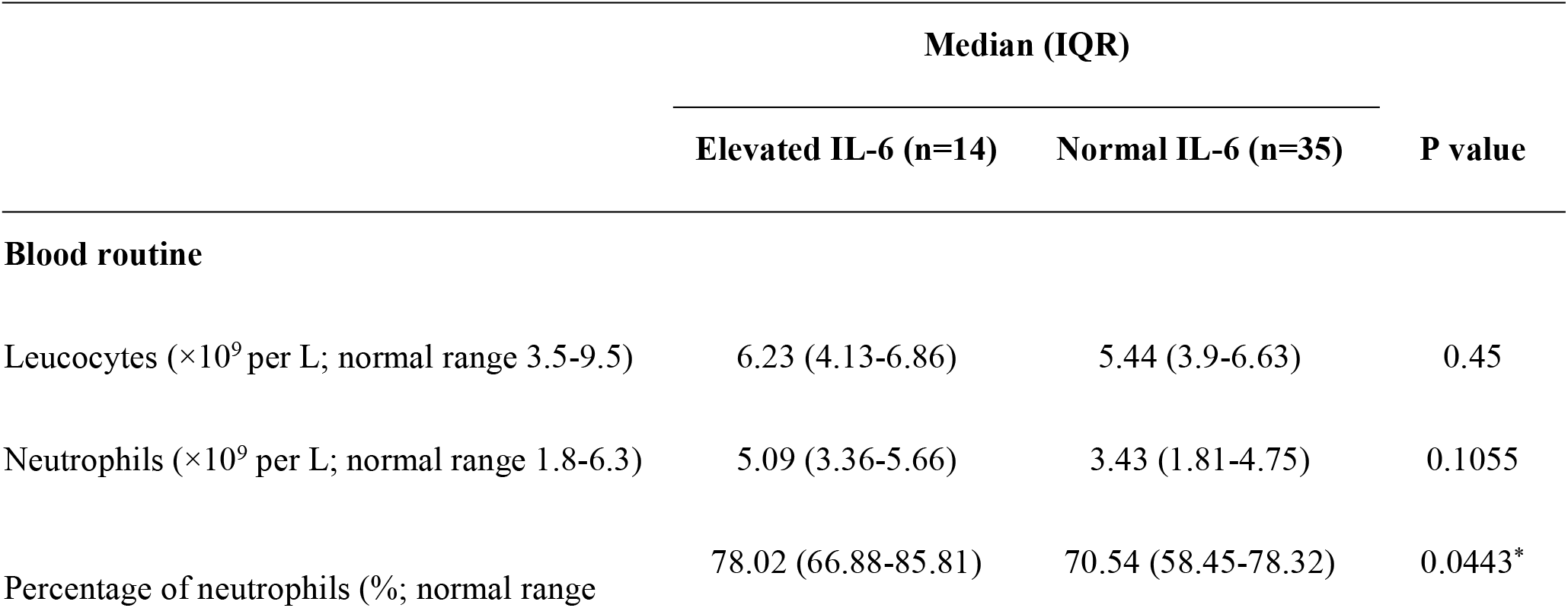
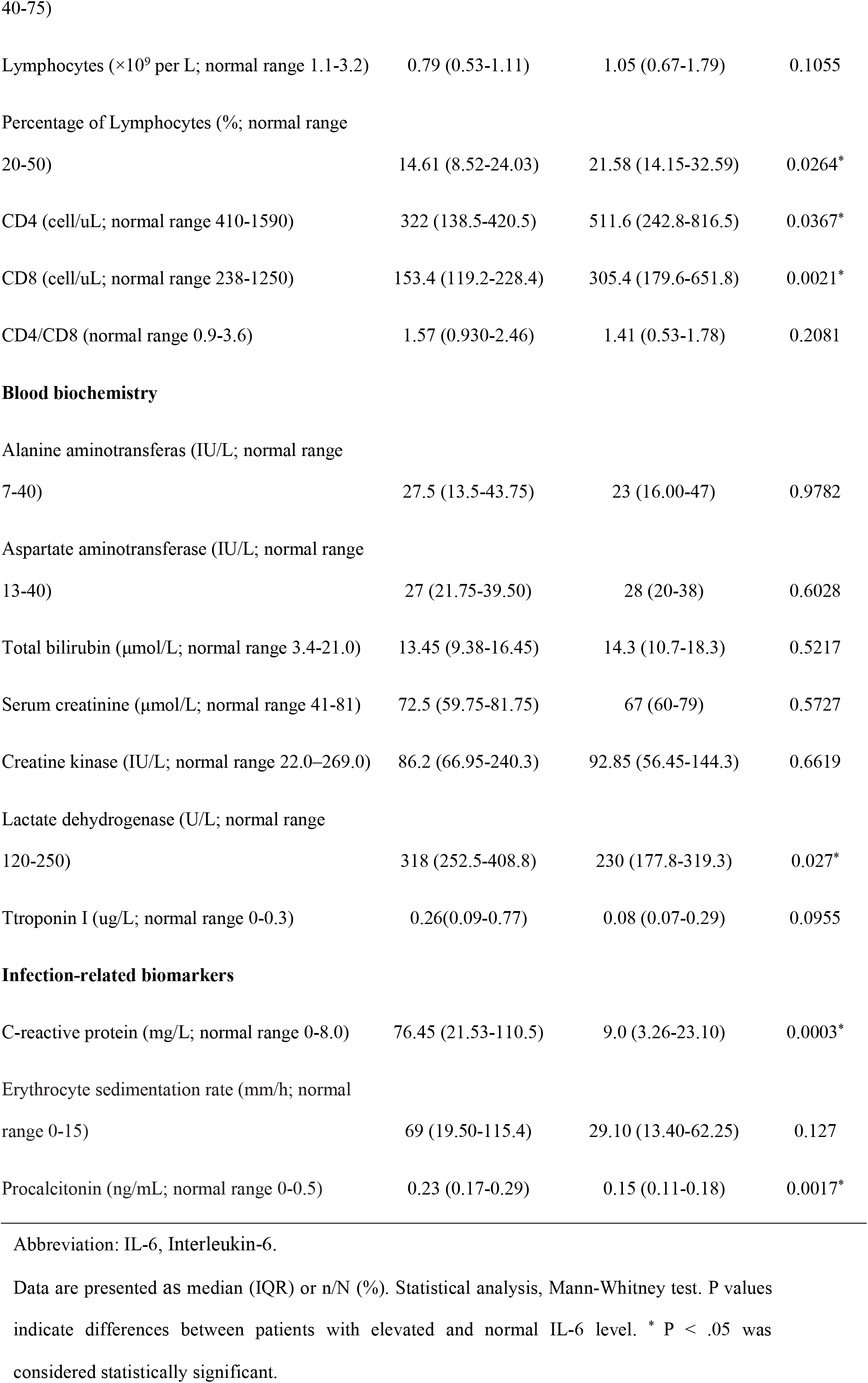
Laboratory findings of patients with elevated and normal IL-6 level.

|  | Median (IQR) |  | P value |
| --- | --- | --- | --- |
|  | Elevated IL-6 (n=14) | Normal IL-6 (n=35) |  |
| **Blood routine** |  |  |  |
| Leucocytes (×10<sup>9</sup> per L; normal range 3.5-9.5) | 6.23 (4.13-6.86) | 5.44 (3.9-6.63) | 0.45 |
| Neutrophils (×10<sup>9</sup> per L; normal range 1.8-6.3) | 5.09 (3.36-5.66) | 3.43 (1.81-4.75) | 0.1055 |
| Percentage of neutrophils (%; normal range | 78.02 (66.88-85.81) | 70.54 (58.45-78.32) | 0.0443* |
| Lymphocytes ($\times 10^9$ per L; normal range 1.1-3.2) | 0.79 (0.53-1.11) | 1.05 (0.67-1.79) | 0.1055 |
| Percentage of Lymphocytes (%; normal range 20-50) | 14.61 (8.52-24.03) | 21.58 (14.15-32.59) | 0.0264* |
| CD4 (cell/uL; normal range 410-1590) | 322 (138.5-420.5) | 511.6 (242.8-816.5) | 0.0367* |
| CD8 (cell/uL; normal range 238-1250) | 153.4 (119.2-228.4) | 305.4 (179.6-651.8) | 0.0021* |
| CD4/CD8 (normal range 0.9-3.6) | 1.57 (0.930-2.46) | 1.41 (0.53-1.78) | 0.2081 |
| Alanine aminotransferase (IU/L; normal range 7-40) | 27.5 (13.5-43.75) | 23 (16.00-47) | 0.9782 |
| Aspartate aminotransferase (IU/L; normal range 13-40) | 27 (21.75-39.50) | 28 (20-38) | 0.6028 |
| Total bilirubin ($\mu\text{mol/L}$; normal range 3.4-21.0) | 13.45 (9.38-16.45) | 14.3 (10.7-18.3) | 0.5217 |
| Serum creatinine ($\mu\text{mol/L}$; normal range 41-81) | 72.5 (59.75-81.75) | 67 (60-79) | 0.5727 |
| Creatine kinase (IU/L; normal range 22.0-269.0) | 86.2 (66.95-240.3) | 92.85 (56.45-144.3) | 0.6619 |
| Lactate dehydrogenase (U/L; normal range 120-250) | 318 (252.5-408.8) | 230 (177.8-319.3) | 0.027* |
| Troponin I ($\mu\text{g/L}$; normal range 0-0.3) | 0.26(0.09-0.77) | 0.08 (0.07-0.29) | 0.0955 |
| Infection-related biomarkers |  |  |  |
| C-reactive protein (mg/L; normal range 0-8.0) | 76.45 (21.53-110.5) | 9.0 (3.26-23.10) | 0.0003* |
| Erythrocyte sedimentation rate (mm/h; normal range 0-15) | 69 (19.50-115.4) | 29.10 (13.40-62.25) | 0.127 |
| Procalcitonin (ng/mL; normal range 0-0.5) | 0.23 (0.17-0.29) | 0.15 (0.11-0.18) | 0.0017* |

Abbreviation: IL-6, Interleukin-6.
Data are presented as median (IQR) or n/N (%). Statistical analysis, Mann-Whitney test. P values indicate differences between patients with elevated and normal IL-6 level. \* $P < .05$ was considered statistically significant.

**Figure 1.**
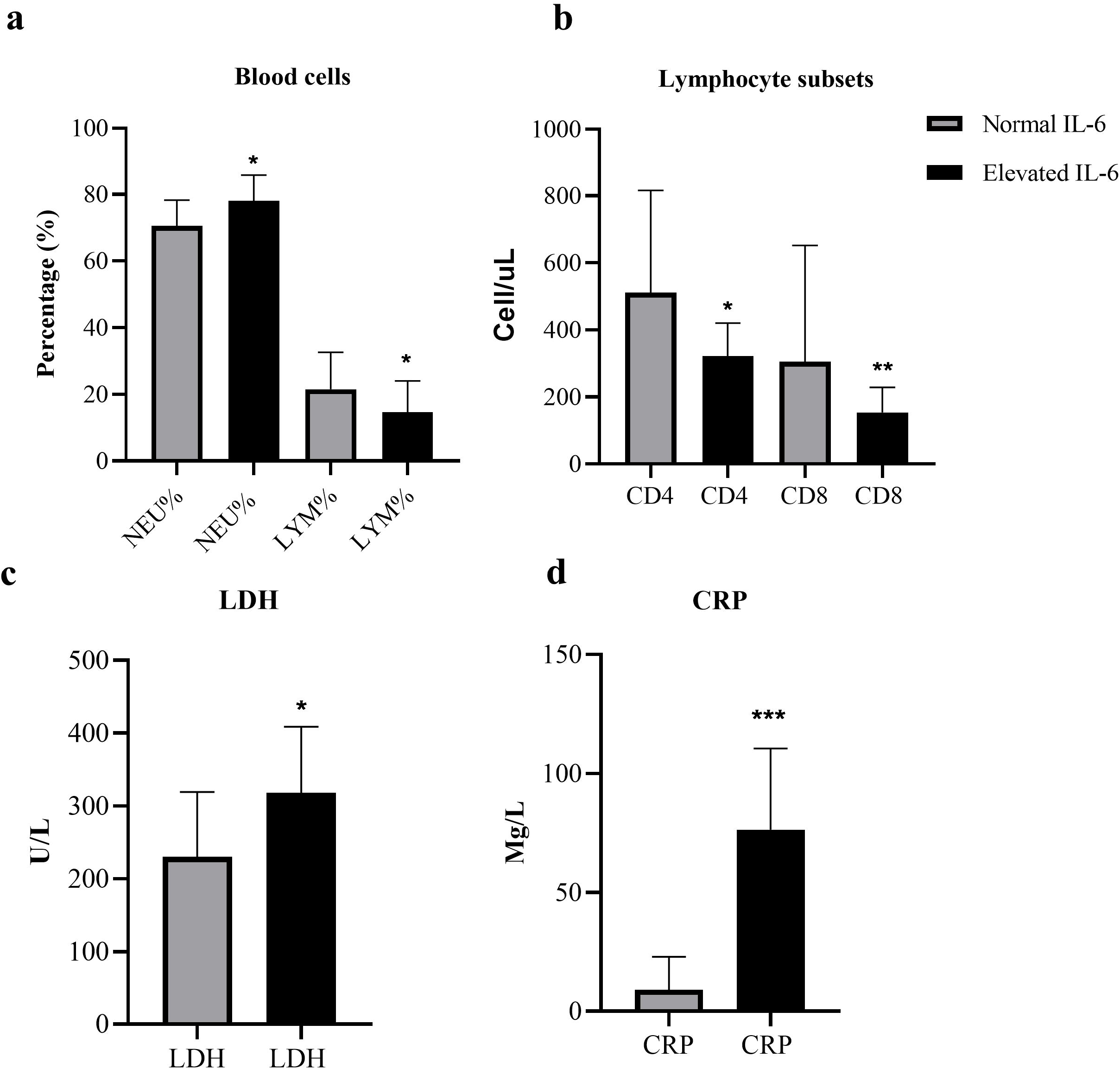
Differences of laboratory findings between patients with elevated and normal IL-6 level. (a) Percentage of NEU and LYM, (b) CD4^+^ and CD8^+^ T cell counts, (c) Detection of LDH levels, and (d) Changes of the infection indicator, CRP in two groups. Data are presented as median (interquartile range, IQR) and analyzed by Mann-Whitney test. All statistical analyses were performed using GraphPad Prism 8.3. P values indicate differences between patients with elevated and normal IL-6 level (^*^ p<.05, ^**^ p<.005, ^***^ p<.001). P <.05 was considered statistically significant. Abbreviations: IL-6, Interleukin-6; lymphocytes percentage, LYM%; neutrophil percentage, NEU%; lactate dehydrogenase, LDH; C-reactive protein, CRP.

## Discussion

This report, to our knowledge, is the first case series of patients with COVID-19 in Anhui Province. As most patients remain hospitalized, we focus on the clinical and laboratory profiles upon their admission. Epidemiological research shows that most patients have been to Wuhan recently. Common symptoms were fever, cough, and chest tightness. However, a significant proportion of patients presented with atypical symptoms such as fatigue, muscle soreness and diarrhea. We also pay attention to patients without fever in which cough may be the only or primary symptom. Therefore, to avoid further transmission, screening and closely monitoring of each suspect remain important. Further studies on the epidemiological characteristics of these atypical cases are recommended.

The most common laboratory abnormalities observed in this study were decreased total lymphocytes, prolonged APTT, elevated LDH, CRP and ESR. Similarities abnormalities between COVID-19 and previously observed betacoronavirus, MERS-CoV and SARS-CoV infection, have been noted [3, 6, 7]. These findings suggest that COVID-19 can cause different degrees of hematological abnormalities and damage of internal organs. The absolute value of lymphocytes was reduced in more than 50% patients. The most significant was the decreased CD4^+^ T cell counts. Previous studies of patients in Wuhan suggested virus invasion could induce a cytokine storm syndrome (CRS) [5, 8]. Of the 14 patients with elevated IL-6, LYM%, CD4^+^ and CD8^+^ T cell counts were significantly decreased and NEU%, CRP and LDH levels increased significantly. Elevated IL-6 may be an important factor leading to T lymphocytes damage and cellular immune deficiency. IL-6 could also be used as an indicator to evaluate infection severity. Therefore, we conclude that IL-6 may be an effective target for prevention or treatment of serve COVID-19 infection. Future large-scale studies are needed to clarify the underlying mechanisms of disease pathogenesis.

COVID-19 belongs to the betacoronavirus. As a single-stranded positive-sense RNA virus, COVID-19 has 79.5% homology with SARS-CoV [9]. Similar to SARS-CoV, angiotensin converting enzyme II (ACE2) is also the cellular entry receptor of COVID-19 [9, 10]. ACE2 is highly expressed in human lung tissue, gastrointestinal tract, vascular endothelial cells and arterial smooth muscle cells [11]. Therefore, all of the organs above may be targets for virus attack. ACE2 effectively hydrolyzes the potent vasoconstrictor angiotensin II to angiotens and is related to hypertension, cardiac function and diabetes [12]. Liu et al. discovered that the Angiotensin II level in the plasma samples increased markedly, suggesting that COVID-19 could induce imbalanced renin-angiotensin system. Drugs of ACE inhibitor (ACEI) and angiotensin receptor blocker (ARB) may be used as potential treatment of COVID-19 infection [13]. As we can see, in patients with underlying diseases, most of them have hypertension. However, no report has focused on the correlation between antihypertensive agents with COVID-19 infection or disease severity. Studies are necessary to evaluate the effectiveness of ACEI and ARB in the future.

Currently, there is no specific therapy for patients with new coronavirus pneumonia. The pathologic mechanisms of disease progression and exacerbation are also unclear. How to relieve the clinical symptoms of critically ill patients, and reduce the severity and mortality of patients still remains challenging. Considering the similarities between SARS-CoV and COVID-19, some pre-clinical drugs against SARS-CoV have been applied to COVID-19 patients. Remdesivir (RDV), a broad-spectrum antiviral nucleotide analogue, is reported to treat MERS-CoV and SARS-CoV infections effectively [14, 15]. A randomized controlled trial was initiated to determine the safety and efficacy of RDV in patients with COVID-19 in Wuhan, China recently. It is crucial to determine host tropism and transmission capacity in terms of prevention of the virus infection [16]. Spike (S) protein mediates membrane fusion through binding with ACE2. Monoclonal antibody against the S protein may efficiently block the virus from entering the host. Convalescent plasma had also been reported to be clinically useful to SARS and MERS patients [17, 18]. If available, convalescent plasma should be used for critically ill patients with COVID-19. However, the appearance of therapeutic plasma requires time and exists only in recovered patients. In our opinion, comprehensive and supportive treatments are essential in the early stage. Additionally, antiviral treatment in early stage and immune activation blockers such as IL-6 blockers, IL-1 blockers in late stage could be tried to control further disease progress leading to ARDS due to excessive immune activation. Targeted treatment should depend on individual differences due to various disease characteristics.

This study has several limitations. First, only 75 patients with confirmed COVID-19 were included. It would be better to include as many patients as possible to get a more comprehensive understanding of COVID-19. Second, more detailed patient information, particularly treatment strategies and clinical outcomes, was unavailable at the time of analysis. Regarding the inflammatory factors, we only measured IL-6 level changes. Future studies should focus on changes of various pro-inflammatory factors, ie IL-1, which may provide precise target treatment options for different patients.

In conclusion, this study provides an early assessment of the clinical and laboratory profiles of COVID-19 patients in Hefei, China. The clinical manifestation of COVID-19 was nonspecific. Specific coronavirus antivirals show proven efficacies in humans are unavailable to date. Antiviral therapy requires a comprehensive and supportive treatment. Targeted therapy should also be determined based on individual clinical manifestations and laboratory indicators.

## Data Availability

The data that support the findings of this study are available from the corresponding author on reasonable request.

## Contributors

ZZ and MY collected the epidemiological and clinical data. JJX contributed to the statistical analysis and drafted the manuscript. YY, TJ, HM, and AZ revised the final manuscript. HH, WL, ZY, XZ, JX, CZ, LL, YL, CD and YQ contributed to clinical and laboratory data acquisition. YG and XM had the idea for the study and take responsibility for the integrity of the data and the accuracy of the data analysis.

## Acknowledgements

We acknowledge all health-care workers involved in the diagnosis and treatment of patients in Hefei. We thank the Chinese National Health Commission for coordinating data collection for patients with COVID-19.

